# Multi-omic approach to identify phenotypic modifiers underlying cerebral demyelination in X-linked adrenoleukodystrophy

**DOI:** 10.1101/2020.03.19.20035063

**Authors:** Phillip A. Richmond, Frans van der Kloet, Frederic M. Vaz, David Lin, Anuli Uzozie, Emma Graham, Michael Kobor, Sara Mostafavi, Perry D. Moerland, Philipp F. Lange, Antoine H. C. van Kampen, Wyeth Wasserman, Marc Engelen, Stephan Kemp, Clara van Karnebeek

**Author notes:** **Correspondence:** Marc Engelen < >, Stephan Kemp < >, Clara van Karnebeek < >. Co-first authors (Phillip A. Richmond, Frans van der Kloet). Co-last authors (Wyeth Wasserman, Marc Engelen, Stephan Kemp, Clara van Karnebeek).

## Abstract

X-linked adrenoleukodystrophy (ALD) is a peroxisomal metabolic disorder with a highly complex clinical presentation. ALD is caused by mutations in the *ABCD1* gene, and is characterized by the accumulation of very long-chain fatty acids in plasma and tissues. Disease-causing mutations are ‘loss of function’ mutations, with no prognostic value with respect to the clinical outcome of an individual. All male patients with ALD develop spinal cord disease and a peripheral neuropathy in adulthood, although age of onset is highly variable. However, the lifetime prevalence to develop progressive white matter lesions, termed cerebral ALD (CALD), is only about 60%. Early identification of transition to CALD is critical since it can be halted by allogeneic hematopoietic stem cell therapy only in an early stage. The primary goal of this study is to identify molecular markers which may be prognostic of cerebral demyelination from a simple blood sample, with the hope that blood-based assays can replace the current protocols for diagnosis. We collected six well-characterized brother pairs affected by ALD and discordant for the presence of CALD and performed multi-omic profiling of blood samples including genome, epigenome, transcriptome, metabolome/lipidome, and proteome profiling. In our analysis we identify discordant genomic alleles present across all families as well as differentially abundant molecular features across the omics technologies. The analysis was focused on univariate modeling to discriminate the two phenotypic groups, but was unable to identify statistically significant candidate molecular markers. Our study highlights the issues caused by a large amount of inter-individual variation, and supports the emerging hypothesis that cerebral demyelination is a complex mix of environmental factors and/or heterogeneous genomic alleles. We confirm previous observations about the role of immune response, specifically auto-immunity and the potential role of PFN1 protein overabundance in CALD in a subset of the families. We envision our methodology as well as dataset has utility to the field for reproducing previous or enabling future modifier investigations.

## 1 Introduction

Adrenoleukodystrophy (ALD) is a rare peroxisomal X-linked degenerative disease (MIM 300100), caused by deficiency of the ABC half-transporter encoded by the *ABCD1* gene. Over 800 different disease-causing loss-of-function ABCD1 mutations have been reported (www.adrenoleukodystrophy.info). Mutations lead to a defect in the import of very long-chain fatty acids (VLCFA) into peroxisomes for further degradation and a subsequent accumulation of VLCFA in plasma and tissues. The overall incidence is 1:17,000. In males, ALD often manifests with adrenocortical insufficiency in childhood (50% before 10 years)(Huffnagel, Laheji, et al. 2019).

During adulthood virtually all male and, eventually, female patients develop a progressive myelopathy termed adrenomyeloneuropathy (Engelen et al. 2014, 2012). Additionally, during childhood or sometimes through adulthood male patients can develop cerebral demyelination, termed cerebral ALD (CALD). It is estimated that eventually more than 60% of male patients develop CALD(de Beer and Scheltens 2016)(Kemp, Berger, and Aubourg 2012). Untreated CALD is often progressive, but can spontaneously arrest in 10 - 20% of patients. It causes vegetative state and death 2-3 years after onset, so early identification as well as careful and frequent monitoring of all male ALD patients is necessary. If diagnosed early, hematopoietic stem cell therapy can be used to halt further progression of cerebral ALD. To ensure timely stem cell therapy for males with CALD, affected individuals are subjected to rigorous neurological and MRI follow-ups that pose considerable physical, emotional and financial burden. As such, the unresolved and unpredictable phenotypic variability of ALD is a crucial roadblock for patient care.

As newborn screening for ALD has recently been implemented, there is an urgent need for identification of markers which may be prognostic of cerebral demyelination in many newly diagnosed patients around the world. Our research focuses on delineating the enormous phenotypic variability in ALD, with the overarching goal of identifying biomarkers prognostic of the advancement to CALD. If successful in identifying biomarkers with prognostic power, then the biomarkers could replace existing expensive monitoring protocols and potentially highlight therapeutic targets, as is the case with other rare genetic disorders. For example, in Spinal Muscular Atrophy, the genes PLS3 and CORO1C were identified as protective modifiers, unravelling impaired endocytosis as a rescue mechanism for the phenotype (Hosseinibarkooie et al. 2016). These modifiers were identified from studies focusing on siblings with discordant disease severity, and are opening novel therapeutic targets for treatment. Patients with ALD may benefit from similar research advances.

Phenotypic discordance in individuals with the same *ABCD1* genotype, including siblings and even monozygotic twins (Korenke et al. 1996), strongly supports the hypothesis that other modifying factors play a role in the progression of the disease (Kemp et al. 2016; Wiesinger, Eichler, and Berger 2015). As yet, however, modifier studies using candidate gene approaches have had little success and resulted in the identification of only a single modifier gene (*CYP4F2*) with limited prognostic power (van Engen et al. 2016). Other candidate variants have been proposed, including a candidate cis-regulatory SNP in the promoter region of *ELOVL1*—a gene involved in VLCFA synthesis (Ofman et al. 2010). The functional consequences of this SNP with respect to the expression of *ELOVL1* in the brain is still under investigation (Kemp, Berger, and Aubourg 2012). The lack of modifier identification could be due to the limited genomic search space that was explored, which to date has focused only on candidate gene approaches. Owing to the small sample size inherent to rare disease cohorts, traditional genome wide association studies (GWAS) approaches are not feasible. Employing a strategy which utilizes family structure may allow for a narrower search space compared to GWAS, while allowing a broader interrogation of the genome than candidate gene approaches. Beyond a search space which involves genetic mapping, other high throughput “omics” technologies allow the exploration of complex biological systems at many levels. It is now possible to identify differences between individuals or phenotypic states at the DNA, methylated DNA, RNA, lipid, and protein levels. Our goal is to delineate personal molecular characteristics that contribute to phenotypic variability in male ALD siblings enabling the identification of biomarkers that prognosticate onset and progression of CALD. Because modifying factors (Génin, Feingold, and Clerget-Darpoux 2008) could also include environmental, epigenetic and microbiome factors (Génin, Feingold, and Clerget-Darpoux 2008; Argmann et al. 2016), multi-omics approaches are key.

In this study, we carefully selected a set of six well characterized brother pairs who have the same *ABCD1* pathogenic allele but are discordant for cerebral ALD: one brother has CALD and the other has no white matter lesions on MRI (non-CALD). The brother pairs are close in age (no more than 2 years apart), and range in age from 6-38 years at sample collection (Table 1). Blood samples were obtained from each of these patients and underwent profiling through five omics technologies (Figure 1) including whole genome sequencing (WGS), RNA sequencing (RNA-seq), EPIC DNA methylation (DNAm) microarray, lipidomic profiling via liquid chromatography mass spectrometry (LCMS), and protein profiling by LCMS. Each omics dataset was processed to quantify/map features, undergo quality control analysis, and then used for group-wise comparisons between CALD and non-CALD phenotype groups using univariate analysis. We first investigated the potential for a single, shared modifier allele which could discriminate the two groups from the WGS data. Next, we systematically compared the groups for each of these omics data sets to find potential markers specific to the phenotype. We aggregated the datasets together after performing pairwise comparisons and identified heterogeneous signals within sub-groups of the 6 families. To the best of our knowledge this is the most comprehensive study to date in terms of systems biology characterization of human ALD using a unique collection of samples.

**Table 1.**
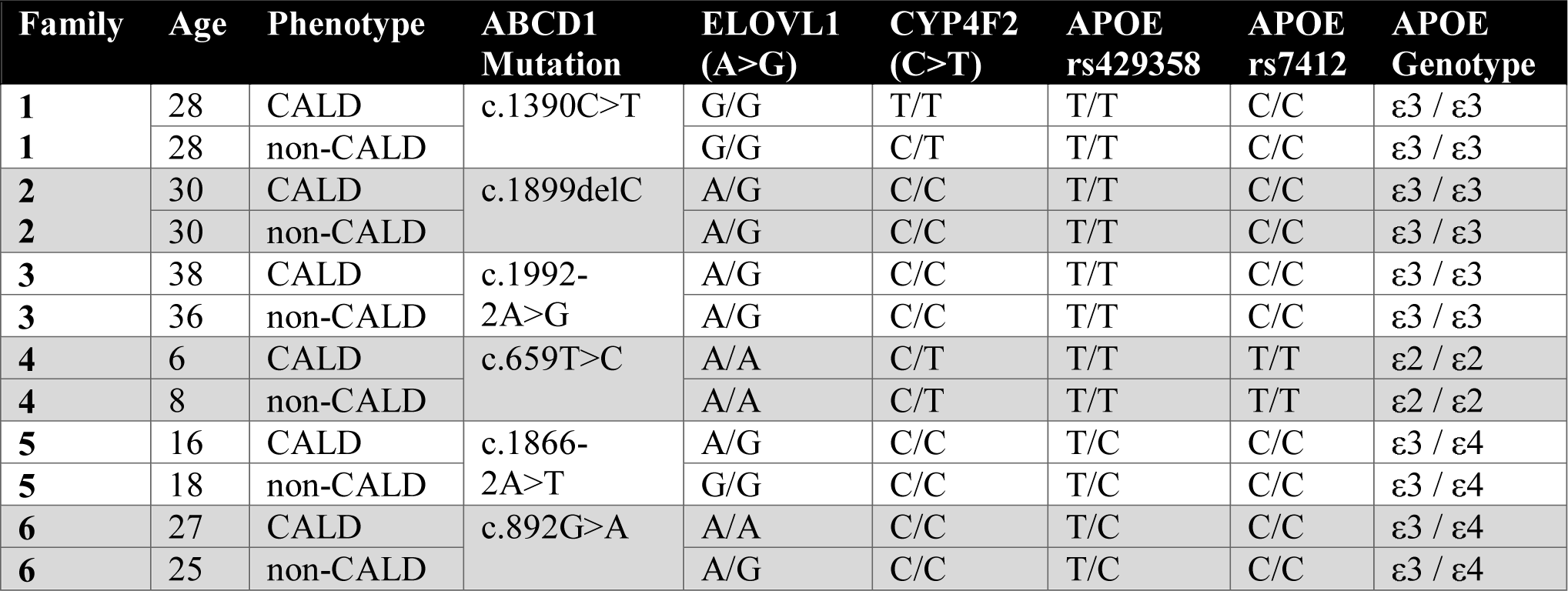
Summary of patients within ALD cohort. The family number, patient ID, age at sample collection, ALD phenotype, *ABCD1* variant, and genotypes for previously associated modifier alleles for all patients within the cohort.

| Family | Age | Phenotype | ABCD1<br>Mutation | ELOVL1<br>(A>G) | CYP4F2<br>(C>T) | APOE<br>rs429358 | APOE<br>rs7412 | APOE<br>Genotype |
| --- | --- | --- | --- | --- | --- | --- | --- | --- |
| 1 | 28 | CALD | c.1390C>T | G/G | T/T | T/T | C/C | ε3 / ε3 |
| 1 | 28 | non-CALD |  | G/G | C/T | T/T | C/C | ε3 / ε3 |
| 2 | 30 | CALD | c.1899delC | A/G | C/C | T/T | C/C | ε3 / ε3 |
| 2 | 30 | non-CALD |  | A/G | C/C | T/T | C/C | ε3 / ε3 |
| 3 | 38 | CALD | c.1992-2A>G | A/G | C/C | T/T | C/C | ε3 / ε3 |
| 3 | 36 | non-CALD |  | A/G | C/C | T/T | C/C | ε3 / ε3 |
| 4 | 6 | CALD | c.659T>C | A/A | C/T | T/T | T/T | ε2 / ε2 |
| 4 | 8 | non-CALD |  | A/A | C/T | T/T | T/T | ε2 / ε2 |
| 5 | 16 | CALD | c.1866-2A>T | A/G | C/C | T/C | C/C | ε3 / ε4 |
| 5 | 18 | non-CALD |  | G/G | C/C | T/C | C/C | ε3 / ε4 |
| 6 | 27 | CALD | c.892G>A | A/A | C/C | T/C | C/C | ε3 / ε4 |
| 6 | 25 | non-CALD |  | A/G | C/C | T/C | C/C | ε3 / ε4 |

**Figure 1.**
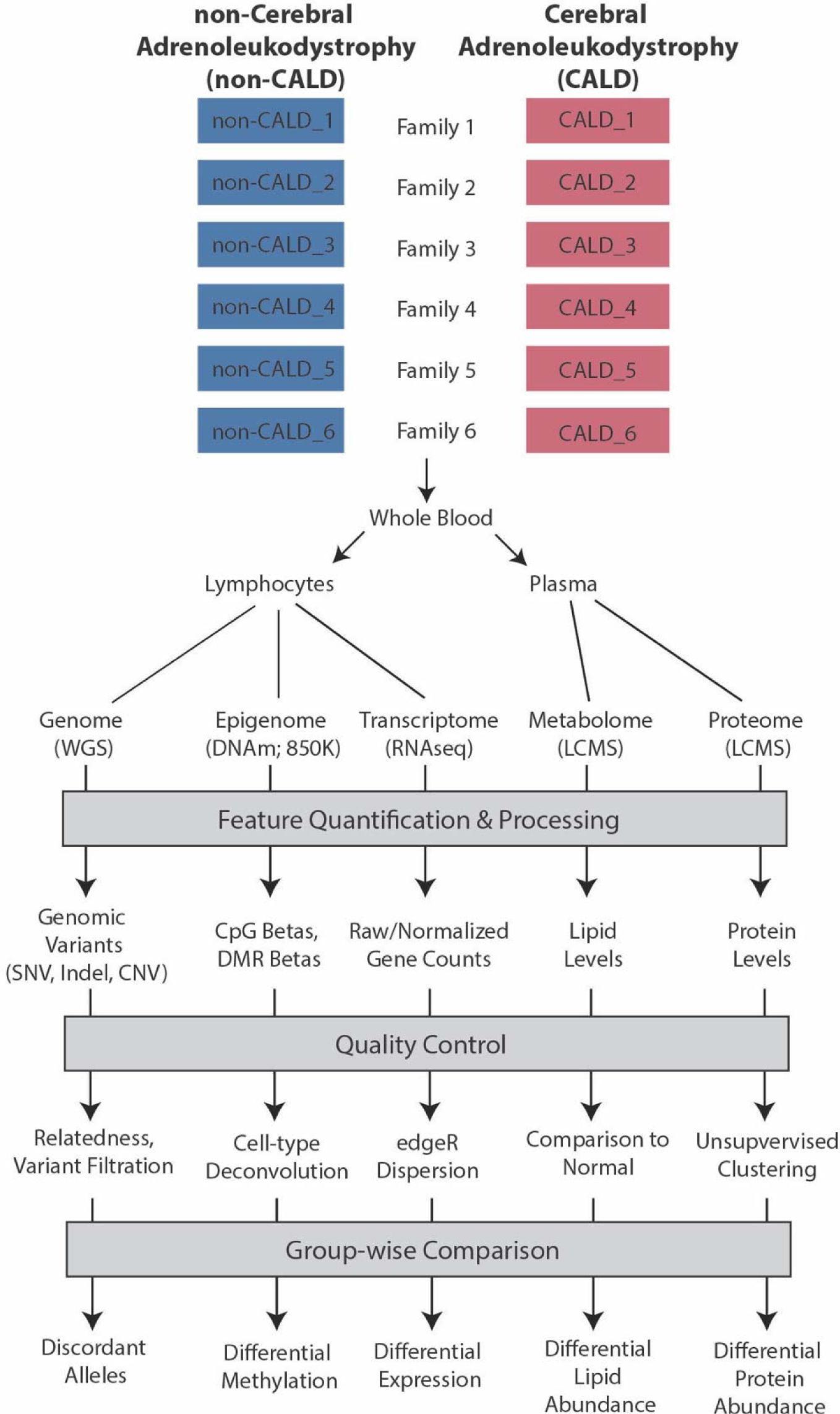
Project overview. An overview of the data and processes involved in the project including samples from two brothers across unrelated families, blood isolated into lymphocytes and plasma, and then profiling with five omics technologies including WGS for the genome, DNA methylation (DNAm) via the 850K EPIC microarray, transcriptome profiling with RNA sequencing (RNA-seq), metabolome profiling with liquid chromatography mass spectrometry (LCMS), and protein profiling with LCMS. These data are then taken through feature quantification/processing, quality control metrics, and group-wise comparison through univariate modeling.

## 2 Materials and Methods

### 2.1 Project Overview

An overview of the project can be found in Figure 1, which depicts the project phases including patient phenotyping/sample collection, multi-omic data collection, feature quantification/processing, quality control, and group-wise comparisons between phenotype groups. In this project, six brother pairs affected by ALD but discordant for the presence of cerebral ALD were included. Patients were selected from the Dutch cohort, an ongoing prospective natural history study (Huffnagel, van Ballegoij, et al. 2019). Blood was drawn from the brother pairs and lymphocyte pellets or plasma was isolated. Lymphocyte pellets were used for whole WGS, RNA-seq, and DNAm. Fasted plasma was used for downstream LCMS analysis identifying either lipid or protein abundances. Data was then processed independently for each of the platforms including feature quantification/mapping, followed by platform specific quality control and group-wise comparisons. Details regarding sample collection, platform specifications, and specific methodology for each analysis performed in this project can be found in the Supplemental Methods section.

### 2.2 Patient selection and phenotyping

All patients were selected from the Dutch cohort, an ongoing prospective cohort study. All patients are examined yearly (by ME) and undergo an MRI of the brain at the time of examination. Samples are collected in the PEROX biobank. The presence of cerebral ALD is defined as the presence of white matter lesions in a distribution consistent with ALD. The classification of the sibs (CALD versus non-CALD) is valid at this time, but the non-affected individuals can theoretically convert to cerebral ALD.

All samples were collected and stored in the PEROX Biobank according to a protocol (METC2015_066) approved by the biobank review board of the Amsterdam UMC (BioBank Toetsingscommissie AMC). All patients provided written informed consent for storage and use of materials for medical research.

### 2.3 Feature quantification and data processing

For each platform, the data was processed independently following best-practices guidelines from the groups generating the datasets. Details regarding feature quantification and assignment at the gene, lipid, protein, and differentially methylated region (DMR) level can be found in the Supplemental Methods section.

### 2.4 Univariate modeling of CALD vs. non cerebral ALD

Using univariate modeling techniques the prognostic power of each lipid, transcript (RNA) or protein is calculated as:

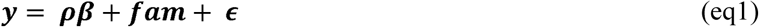

in which y is the observed value and *ρ* the phenotype (0 or 1), *β*is the weight and fam the family cofactor. *ε* is the remaining error. Because methylation of DNA changes with age (McEwen et al. 2018), age is also included as a cofactor:

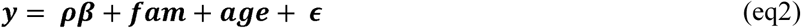

when analysing the DNAm results. The significance (p-value) of the discriminating phenotype (fixed) and family (random) effects are determined by ordinary least squares modelling (OLS) of the data using the model from Eq. 1 in case of lipid and proteomic data (Harrison et al. 2018). In the case of methylation and RNA sequencing data the p-values are determined by maximum likelihood estimates (MLE) of the fixed and random effects using Limma (Ritchie et al. 2015) and edgeR respectively (Ritchie et al. 2015; Robinson, McCarthy, and Smyth 2010).

### 2.5 Allele comparisons in whole genome sequencing data

Details on data processing, including variant calling and comparing across samples can be found in the Supplemental Methods section. Briefly, allele comparisons were performed in whole genome sequencing data on jointly genotyped variant datasets. For SNVs and indels, variants were jointly genotyped and converted into a GEMINI database (Paila et al. 2013). This database was then queried to identify subsets of discriminating alleles. For structural variants and mobile element insertions, custom scripts were used to identify discordant genotypes from annotated jointly genotyped variant tables. Discordant genotypes, stored as unique variant identifiers, were then placed into Intervene for intersection analysis (Khan and Mathelier 2017).

### 2.6 Aggregation of signal across platforms

To assess the added value of combining the different platforms, significant signals prior to multiple testing correction were collected for each omics platform and intersected at the annotated gene level (hg19). Because of the lack of a clear mapping of lipids to genes the lipidomics platform was excluded from this intersection allowing 4 possible intersections; DNAm-RNA, DNAm-Protein, RNA-Protein and the overall intersection of DNAm-RNA-Protein. Further investigation of a shared signal was performed by clustering the first 3 principal components (i.e. capturing the most variance) of the log fold changes (top 10 and p<0.05) of the combined platform data (including lipids).

### 2.7 Assessing contribution of family effect per feature

The data were modelled using the equations (above) in which both phenotypic and family effects are estimated. We partitioned the variance for the lipids, proteins, and RNA datasets to identify the contribution of the family effect, the phenotype effect, or the residual variance using the variancePartition package (Hoffman and Schadt 2016). The same was repeated for the DNA methylation dataset with the addition of the age, and phenotype:age variance terms. Next, we plotted the top two principal components for each omics dataset before and after the removal of the variance contributed from the family effect with the limma:removeBatchEffect tool (Ritchie et al. 2015). Lastly, to determine the sensitivity/specificity of the findings for leaving out a one or two families all the analyses (excluding methylation data analysis) that were run for the case of all families were repeated with a one or two families left out (e.g. without fam 1, without fam 2, without fam 1 and fam 2, etc.). We encapsulated this information in separate upset plots for each platform.

## 3 Results

### 3.1 Lipidomics analysis of a fatty acid storage disorder

Patients affected by ALD have a buildup of VLCFAs within cells in the body. Recent mass spectrometry advances allow for broad, untargeted profiling of lipids (Huffnagel, Dijkgraaf, et al. 2019). We applied LCMS from plasma samples of each of the patients within this cohort as well as matched control samples.

First, we identified differential lipid abundances between ALD (both non-CALD and CALD) samples and control samples, with 139 lipids passing the threshold of p-value < 0.05 (Eq. 1, OLS), and 17 lipids remaining significant after multiple testing correction (Bonferroni) (Figure S1, Table S1) (Methods). The measured lipids are plotted as a volcano plot, that is the log2 fold change of ALD over control versus corrected p-value (Figure 2A). We confirm that untargeted lipidomic profiling can distinguish ALD from control samples via principal component analysis, and also capture the expected differentially abundant lipids between control and ALD samples including the known ALD biomarker LPC(26:0) (Figure 2B,C).

**Figure 2.**
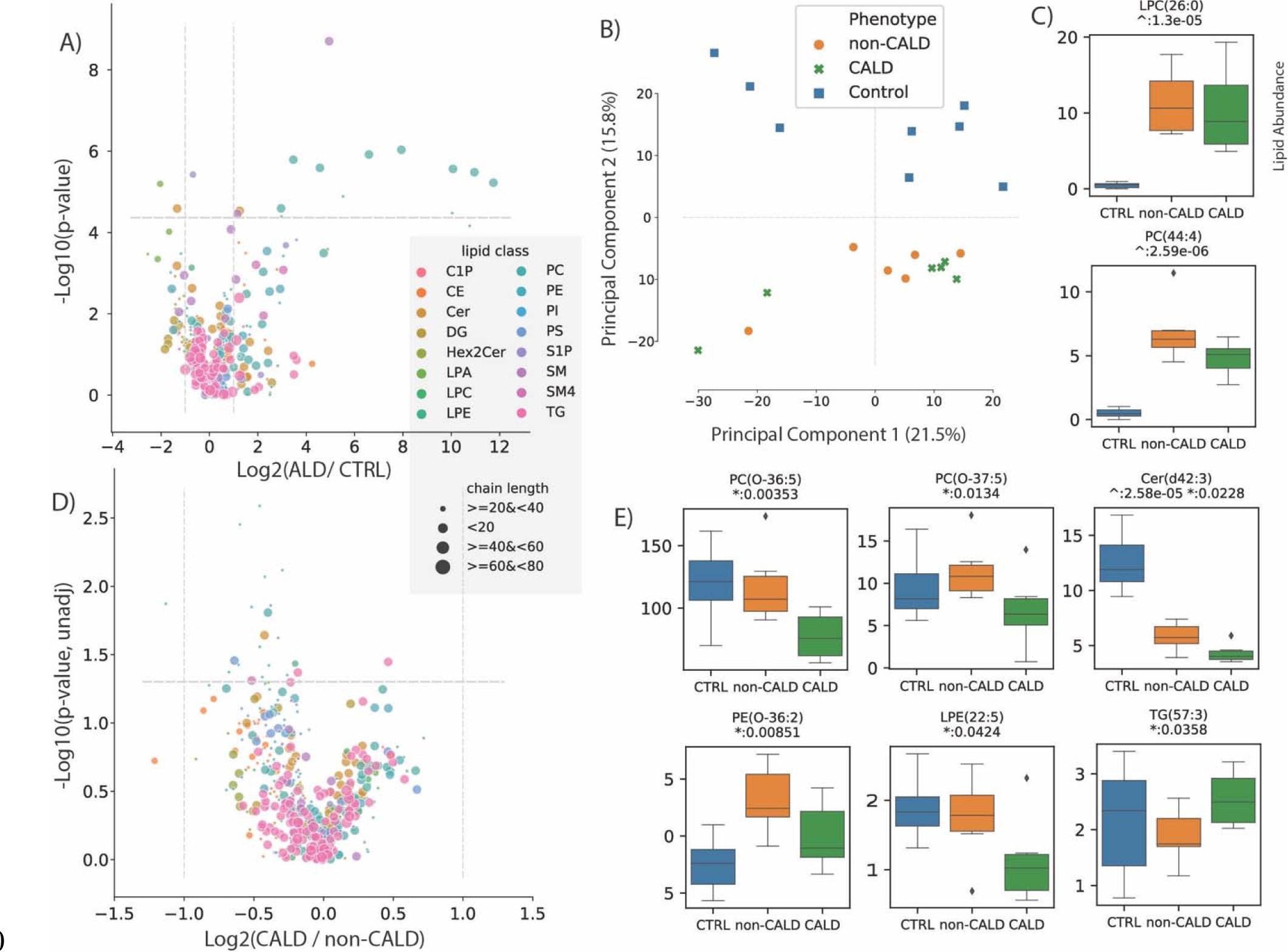
Lipidomic analysis of ALD. The univariate analysis comparing the lipid abundances between control vs ALD, and CALD vs non-CALD is depicted. A) Volcano plot showing the log2 fold change between ALD and control (CRTL) samples for all of the measured metabolites within the LCMS assay, versus the −log10 transformed adjusted p-value. B) Principal component analysis plot showing the first two principal components which can discriminate between control (blue) and ALD (orange:non-CALD, green:CALD) samples. C) Boxplots showing the abundances of a known marker for ALD, LPC(26:0), and another lipid differentially abundant between ALD and control samples. Values are lipid abundances measured on LCMS. D) Volcano plot showing the log2 fold change between CALD and non-CALD samples versus the −log10 transformed p-value. E) Boxplots for lipids different between CALD and non-CALD before p-value correction. For A) and D), the lipids are coloured according to their assigned class and their size corresponds to the lipid chain length. For boxplots: ^ represents unadjusted p-values of comparison between ALD and control, * represents unadjusted p-values of comparison between CALD and non-CALD.

Next, we compared CALD and non-CALD groups for differences in lipid abundance which could act as markers of cerebral demyelination. Of note, the principal component analysis which separates ALD from control did not separate CALD from non-CALD, i.e. the differences in lipid profiles between these two phenotypes are much less pronounced than the differences separating ALD patients from controls (Figure 2B). The measured lipids are plotted as a volcano plot, that is the log2 fold change of CALD over non-CALD versus transformed p-value (Figure 2D). In total 22 lipids were found to have different abundances between the two groups with p-value <0.05, however none of the lipids remained significant after correcting for multiple testing (Table S2). The observed differences are much smaller between CALD and non-CALD compared to ALD and control, as highlighted by the differences in fold change axes (Figure 2A, D). Interestingly, there was a higher abundance in the non-CALD group for several key VLCFAs involved in ALD including PC(44:4) and Cer(d42:3), the latter reaching p-value < 0.05 (Figure 2C,E). While some lipids show a relatively large fold change between CALD and non-CALD groups as a whole, the signal is not consistent for every family. An example of this can be seen in SM(d36:2) or PS(43:3) (Figure S2). This limits the prognostic power of these lipids as consistent markers delineating the phenotype. Lastly, we observed a large range of lipid abundances within the control group for several of the differential lipids between CALD and non-CALD, which could indicate that these lipids are variable within healthy individuals and the signal we observe between CALD and non-CALD could be due to noise or variation in the healthy population (Figure 2E, Figure S1).

### 3.2 Discordant genotype analysis for the identification of a modifier allele

Using whole genome sequencing, we investigated a range of variant classes for discordant alleles between siblings. These discordant alleles are then intersected across multiple families under the hypothesis that polymorphic differences contribute to cerebral demyelination.

We first focused on alleles which emerged from previous modifier studies to see if they are confirmed. Proposed modifier alleles from target gene studies have identified two candidates within ELOVL1 (rs839765) and CYP4F2 (rs2108622) (van Engen et al. 2016; Kemp et al. 2012). Within this cohort, those modifier alleles do not segregate with ALD phenotype (Table 1), nor are the genotypes shared or lacking in the confidently phenotyped CALD patients. Furthermore, it has been suggested that APOE genotypes--which are a combination between two SNP sites to produce APOE2 (ε2), APOE3 (ε3), and APOE4(ε4) alleles--may be markers of disease severity and cerebral progression (Orchard, Markowski, et al. 2019). These APOE alleles do not segregate with disease nor are they shared by all CALD patients. Together, these results suggest limited prognostic power of these alleles, and perhaps supports heterogeneous contributions of genetic background to disease progression.

Next, for several variant classes, we performed a discordant analysis between siblings and intersected these alleles across families (Figure 3). We considered four genotypic categories termed dominant protective, recessive protective, dominant damaging, or recessive damaging based on the genotype (heterozygous: dominant or homozygous: recessive) and the sibling which carries the genotype (CALD: damaging or non-CALD: protective). Performing this genotypic analysis on SNVs and indels, we identified ∼6.0×10^5^ discordant candidate variants in the dominant categories from each family, and ∼3.0×10^5^ discordant candidate variants from the recessive categories (Figure 3A, Table S3). Despite the large number of discordant candidates per family, intersecting these sets across families reduces the candidates dramatically, resulting in only two candidate variants at the intersection of all 6 families (Figure 3B) (Table S4). A recessive damaging variant downstream of the *WIBG*/*PYM1* gene (rs7980776) and recessive protective allele (rs55639747/rs61327784) within a *CCDC67*/*DEUP1* intronic region (Figure S3 & S4). We validated our approach with a parallel pipeline utilizing the new DeepVariant tool (Poplin et al. 2018), which claims higher accuracy than GATK HaplotypeCaller (Table S4). There is high concordance between the two variant call sets, and they produced the same two variants within the intersection. A single additional variant was reported using DeepVariant under the recessive damaging model, however the variant did not pass the manual inspection quality assessment. In silico analysis of both variants suggests these variants have little functional effect, and the associated genes did not link to the cerebral demyelination phenotype (Supplemental Results).

**Figure 3.**
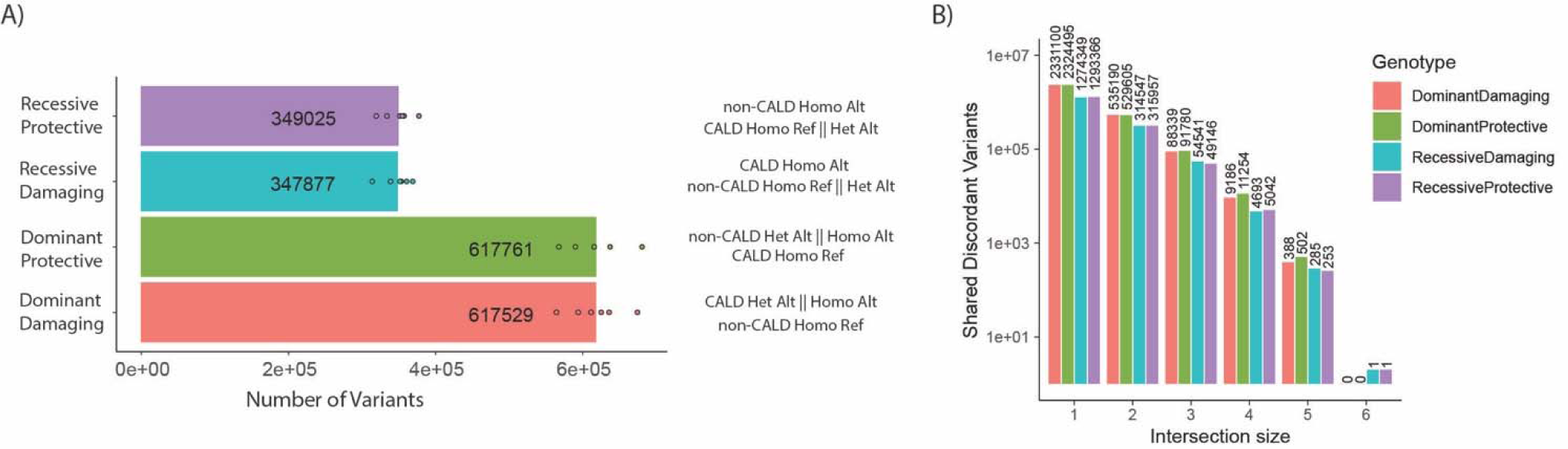
Discordant genotype analysis. A) Number of discordant genotypes in each category for each of the 6 families, with description of genotypes for non-CALD and CALD pairs per genotypic category (per-category means are displayed). B) Upon intersection of discordant genotypes, the number of variants which exist within any intersection with set sizes of 1-6, meaning the set size of 6 is the intersection of all families, and a set of 1 are discordant variants only found in one family.

For the other variant classes, including structural variants (SVs) and mobile element insertions (MEIs), we performed joint genotyping to identify shared and discordant alleles in the same manner as SNVs and indels. We identified ∼1500 and ∼400 SVs in the dominant and recessive categories respectively, and ∼400 and ∼100 MEIs (Table S3). Unsurprisingly, these discordant events were not shared across more than 4 families (Table S4). We further manually inspected the regions around the discordant SNVs/indels identified above, and did not find any other segregating SVs or MEIs.

Lastly, we extended our discordance analysis to the mitochondrial genome to examine candidate alleles which may show evidence of heteroplasmy which are not shared between two siblings. We identified that between 129 and 547 mitochondrial variants per sample, of which 52 to 476 are heteroplasmic, and none are consistent discriminating variants between phenotypes shared across all families (Table S5). Further, if we aggregated at the gene level, we did not find any heteroplasmic variants consistent across the same gene.

In recognition of a study limitation--the fact that some non-CALD patients may progress to CALD--we further intersected alleles shared by all CALD patients. These variants were annotated by impact or as eQTLs defined in GTEx (Supplemental Methods). There were 48 variants present in the heterozygous state across all CALD patients, where no non-CALD patients were heterozygous (Table S6). Of these, a haploblock containing 20 variants was identified overlapping the gene *TPCN2*, including a missense variant (rs3750965) (Figure S5). Interestingly, the only patient which was homozygous for this variant is the youngest non-CALD patient within the cohort, suggesting that this gene could be of significance should the patient develop the cerebral demyelination phenotype.

### 3.3 Univariate modeling of phenotype differences across omics platforms

Beyond identifying a single genetic modifier allele, the omics platforms allow for the identification of candidate molecular signatures which can discriminate between the CALD and non-CALD phenotypes. Using univariate analysis we identify differences across each platform at the feature level, to search for a signal which can be used as a marker for transition to CALD. Further, we leverage these molecular signatures to provide insight into the pathogenesis of cerebral demyelination.

#### 3.3.1 Transcriptomics

Examining RNA expression using RNA-seq provides a measurement for nearly all expressed protein coding genes in the genome. Differential gene expression was calculated between the two phenotype groups using the univariate model accounting for family effect (equation 1). There were 199 genes found with a p-value < 0.05, although none remained significant after multiple testing correction (Bonferroni) (Figure 4A,B, Figure S6). This is likely due to the low number of samples and relatively small differences that were observed between the two groups. Furthermore, many of the genes identified as significant were inconsistent in one or more of the sibling pairs, limiting the diagnostic utility as a marker (Figure 4B). Despite not having significant genes after multiple testing correction, we performed enrichment analysis using GO (gene annotation) and KEGG (pathway annotation) to derive insights based on the 199 genes passing a threshold of p-value < 0.05 (Figure S7). Of note, elevated interferon related processes suggest that the host may be reacting to pathogens activating the immune system (Hoffmann, Schneider, and Rice 2015). It is therefore no surprise that 3 chemokines (*CXCL6, CXCL8* and *IFI27*) were found in the top 10 differentially expressed genes. Amongst the remainder of the proteins encoded by the top 10 differentially expressed genes, the D-Xylulokinase gene (*XYLB*) encodes for the protein that catalyzes the ATP-dependent phosphorylation of D-xylulose to produce xylulose-5-phosphate (Xu5P) therefore *XYLB* may play an important role in metabolic disease given that Xu5P is a key regulator of glucose metabolism and lipogenesis (Bunker et al. 2013). *GATM* has been associated with statin intolerance (V Willrich et al. 2018) and its function to catalyze creatine and possibly affect the production of ceramides (Turer et al. 2017).

**Figure 4.**
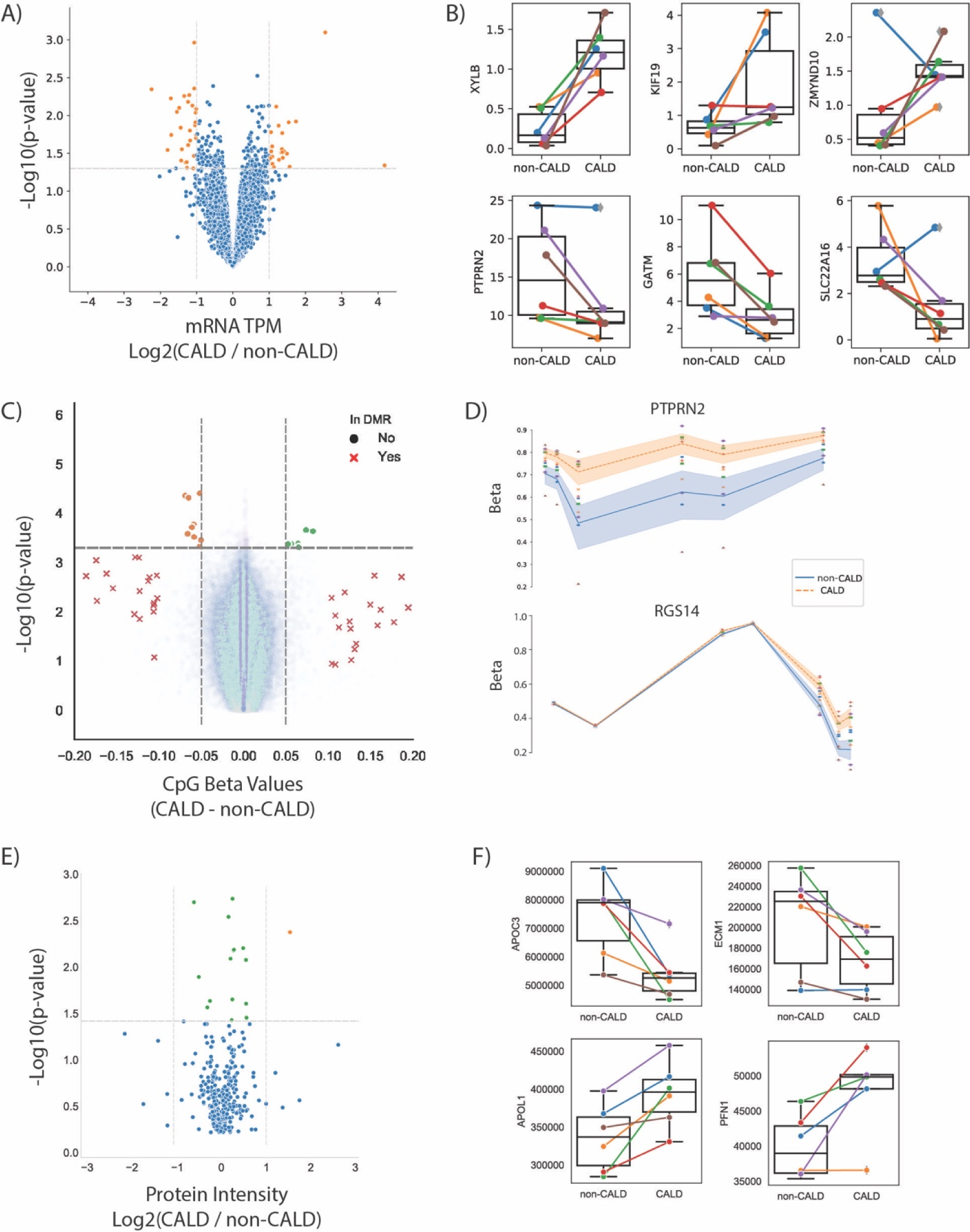
Multi-omic analysis. A) Volcano plot showing p-value and log2 fold change of gene expression from RNA-seq. Significant genes at p < 0.05 (orange dots), non-significant genes (blue dots). B) Selected genes plotted as normalized RNA-seq values with boxplots for each group where each line/point is coloured by family. C) Volcano plot of DNA methylation over CpG probes from EPIC array, with non-significant (p > 0.05) DMR probes (blue dots), significant CpGs at the DMR level (red Xs), higher methylated non-CALD probes (orange dots), and higher methylated CALD probes (green dots). D) DNAm over two significant DMRs within *PTPRN2* and *RGS14*, points coloured by family and lines coloured by phenotype, with shading denoting inner quartile range. E) Volcano plot of protein levels from LCMS with non significant proteins (blue), significant proteins with log2 fold-change (CALD / non-CALD) of −1 to 1 (green), and log2 fold-change greater than 1 (orange). F) Selected proteins which passed the p-value threshold of 0.05.

*MYOB1B* is a protein that may participate in a process critical to neuronal development and function such as cell migration, neurite outgrowth and vesicular transport (Sittaramane and Chandrasekhar 2008).

#### 3.3.2 Epigenomics

DNA methylation has been linked to changes in gene expression, and is an important readout of some environmental impacts upon the cell. Measuring DNA methylation is typically done at specific methylation sites (CpGs), and then aggregated across regions where several sites have similar trends of methylation levels to find differentially methylated regions (DMRs). Here, we used the MethylationEPIC BeadChip which targets over 850,000 CpGs. Using LIMMA modeling including age as cofactor (equation 2) 264 CpGs had a nominal p-value <0.0005. Of these 264 CpGs, 16 passed the delta beta (i.e. difference between methylation levels of CALD vs non-CALD) of >5% (Table S8). When aggregating these loci into a DMR analysis, we identified 22 regions passing thresholds of FDR<0.05 and >10% methylation change (Figure 4C). Multiple CpGs map to the same gene and show a large delta beta, which we identified in the genes *PTPRN2* and *RGS14* (Figure 4D). *RGS14* may alter calcium levels to enhance long term potentiation and learning (Lee et al. 2010).

Due to its presence in neurosecretory vesicles, *PTPRN2* has been implicated in insulin and neurotransmitter exocytosis (Sengelaub et al. 2016). Furthermore, *PTPRN2* hypermethylation has been identified within a separate study which compared DNA methylation between CALD and non-CALD patients (Schlüter et al. 2018).

#### 3.3.3 Proteomics

In addition to profiling lipids, LCMS can be used for high throughput profiling of proteins thus enabling the identification of differential protein abundances between samples. Applying proteomics to these 12 patients yielded a quantification of 5,862 peptides which were matched against 351 protein groups. Comparing CALD and non-CALD groups, we found 16 proteins with differential abundances (p < 0.05) (Figure 4E,F; Figure S8; Table S9). Investigating the top hits we find 4/16 proteins associated with immunoglobulin heavy chain (IGHV4-34, IGHV3-30, IGHV3-7, and P0DOX6), 2/16 are associated with immunoglobulin kappa variables (IGKV6D-21 and IGKV1D-33), and with P0DOX8 also being related to immunoglobulin, half of these proteins are related to the immune system (Parra et al. 2016). All of these immunoglobulin proteins were up-regulated in the CALD samples. Also related to the immune system is CD5L, a secreted glycoprotein that participates in host response to bacterial infection (Sanjurjo et al. 2015) and is also known to regulate lipid biosynthesis (Wang et al. 2015). Beyond immune system proteins, we identified proteins associated with the brain or with involvement in lipid metabolism. ECM1 has been associated with lipoid proteinosis in which brain damage develops over time and is associated with the development of cognitive disabilities and epileptic seizures (Zhang et al. 2014). The role of APOL1 is not yet clear but it has been associated with the lipid biology in the podocyte (Fornoni, Merscher, and Kopp 2014). Copy number variants of MINPP1 have been associated with varying IP6 levels (Waugh 2016) and IP6 has been reported to suppress lipid peroxidation (Foster et al. 2017). APOC3 is a key player in triglyceride-rich lipoprotein metabolism (Ramms and Gordts 2018) and regulated by the peroxisome proliferator-activated receptor-α (Liu et al. 2015). PFN1 has recently been reported in a CALD study which looked at markers of autoreactivity, identifying anti-PFN1 antibodies present in a large proportion of CALD patients (Orchard, Nascene, et al. 2019). Together, these protein signals could have significance with respect to the pathophysiology of cerebral demyelination, by highlighting differences around proteins involved in lipid metabolism as well as immune response.

#### 3.3.4 Estimating variance of family effect

The univariate modeling of CALD vs. non-CALD for each of the individual omics platforms was unsuccessful in identifying significant hits after multiple testing correction. While traditional multiple testing correction methods may be too strict for the omics technologies, we still cannot rule out the possibility that our top hits arise by chance due to variability. Furthermore, our top hits per platform still exhibited a high amount of variance between families, and a lack of consistent signal in molecular features across the entire cohort (Figure 4 B, D, F). Within our model we included the effect of the family on the level of the measured signal, and thus we are able to capture the contribution of family structure to a feature’s abundance (equation 1, equation 2). To illustrate the contribution of these effects, we partitioned the variance contribution within our linear models (Methods). The phenotype effect, total family effect, and residual variance were extracted from our model for each of the features within the RNA-seq, proteomics, and lipidomics platforms (Figure S9). As DNAm varies with age we additionally extracted the variance contributed from the age or phenotype-by-age effects. Clearly, the contribution of variance from the phenotype is small in the majority of features across all omics datasets, and a large residual variance indicates a high level of noise present in these high dimensional assays (Figure S9). We further demonstrated the heterogeneity in the data by subsetting the families and then repeating comparisons between CALD and non-CALD phenotypes. By leaving out one or two families, the *β*in equations 1-2 are re-evaluated for the RNA, protein, and lipid datasets. The number of candidates increased with removal of each family, which could be interpreted as potential modifier signatures present in a subset of families, but absent from others (Figure S10).

### 3.4 Integrating multi-omic datasets

As it was our intention to identify molecular marker features underlying cerebral demyelination, we investigated the omics datasets independently to identify a consistent signal. However, owing to a large amount of inter-family variance, we are limited in our ability to identify a statistically significant feature which separates the two phenotypes. As the multi-omic assays should be complementary to each other, we searched for genes which showed differences between the groups in multiple assays. We searched the phenotype comparison between all families, as well as the results from the leave-one-out analysis, wherein we withheld a family and repeated the modeling between the two phenotype groups (Methods). Intersections showed overlapping evidence at the DNA methylation and RNA levels, as well as overlap between RNA and protein levels, for eight genes. Focusing only on the intersection of all families, only *PTPRN2* has differential signal from both DNA methylation and RNA levels (Figure 4 B,D). Additional genes were identified in the leave-one-out subsets (Table 2).

**Table 2.** Intersections of significant hits from multiple platforms. For each comparison including all families, and each possible 5×5 comparison between CALD and non-CALD, the significant hits (p-value < 0.05 before multiple testing correction) from DNA methylation (DNAm), RNA-sequencing (RNA), and protein LCMS (Protein) were intersected. (↑ means up-regulated/higher for CALD, ↓ means lower in CALD).

| Comparison | DNAm & RNA | DNAm & Protein | RNA & Protein | DNAm & RNA &<br>Protein |
| --- | --- | --- | --- | --- |
| all_families | PTPRN2(↑ - ↓) | □ | □ | □ |
| wo_fam_1 | □ | □ | □ | □ |
| wo_fam_2 | □ | □ | □ | □ |
| wo_fam_3 | HLA-DQB1(↓-↓),<br>IL5RA(↓ - ↑),KIF19(↑- ↓) | □ | □ | □ |
| wo_fam_4 | □ | □ | □ | □ |
| wo_fam_5 | □ | □ | ICAM1(↑ - ↓), APOL1(↑ -<br>↑), CD14(↑ - ↑) | □ |
| wo_fam_6 | □ | □ | JCHAIN(↑ - ↑) | □ |

In the multi-omic data we observed that several of the molecular features have trends of differential abundance/expression in a subset of the families. To illustrate this, and attempt to identify clusters within the data, we gathered per-family log2-fold-change of CALD over non-CALD for the top hits from the lipid, protein, and RNA datasets. We took this approach because it removes the differences in absolute levels of expression between families. Noticeably, the fold-change values are not consistent for each family, as evidenced by a lack of consistent colouring for each of the features (rows) within the heatmap (Figure S11 A). Family 2 and family 6 were more similar in their CALD/non-CALD ratios for these features. This is further supported by a principal component analysis, wherein family 2 and family 6 are separated from the other four families on the first principal component (Figure S11 C). However, this trend does not hold when the set of features is increased to all hits with p-value < 0.05 across the three platforms, as family 1 and 5 cluster together with the other four families as an outer group (Figure S11 B). Thus, clustering these families based on top differential features does not reveal confident sub-groupings within the small cohort.

### 3.5 Specific modifier hypothesis testing

Finding molecular markers which delineate cerebral demyelination in patients with ALD is an ongoing research problem. Additionally, understanding the pathophysiology of cerebral demyelination and potential disruption of the blood brain barrier has implications for diseases beyond ALD. Different hypotheses have been suggested, including involvement of the immune system in autoreactivity or as a response to severe viral infections. Using the multi-omics dataset, which gives us insight into the complexities of the underlying complex biological system, we tested recently proposed modifiers of cerebral demyelination to see if there is evidence of their discriminatory power within the blood samples profiled in our dataset.

It has recently been demonstrated that autoreactivity to profilin (PFN1) occurs in patients affected by CALD, and may be a discriminating marker of cerebral demyelination (Orchard, Nascene, et al. 2019). We investigated differences in PFN1 methylation, RNA, and protein levels between CALD and non-CALD patients to see if this observation is confirmed in our dataset. At the methylation and RNA level, we did not see a consistent signal differentiating the CALD and non-CALD groups, but at the protein level we observe an increased amount of PFN1 in the CALD group for four out of six families (Figure 5 A, B, Figure 4F). This is consistent with the observation from the previous study that not all patients exhibit PFN1 autoreactivity, and the dramatically increased protein levels could precede or act as biomarkers of the autoimmune response within the subset of patients who exhibit this trend.

**Figure 5.**
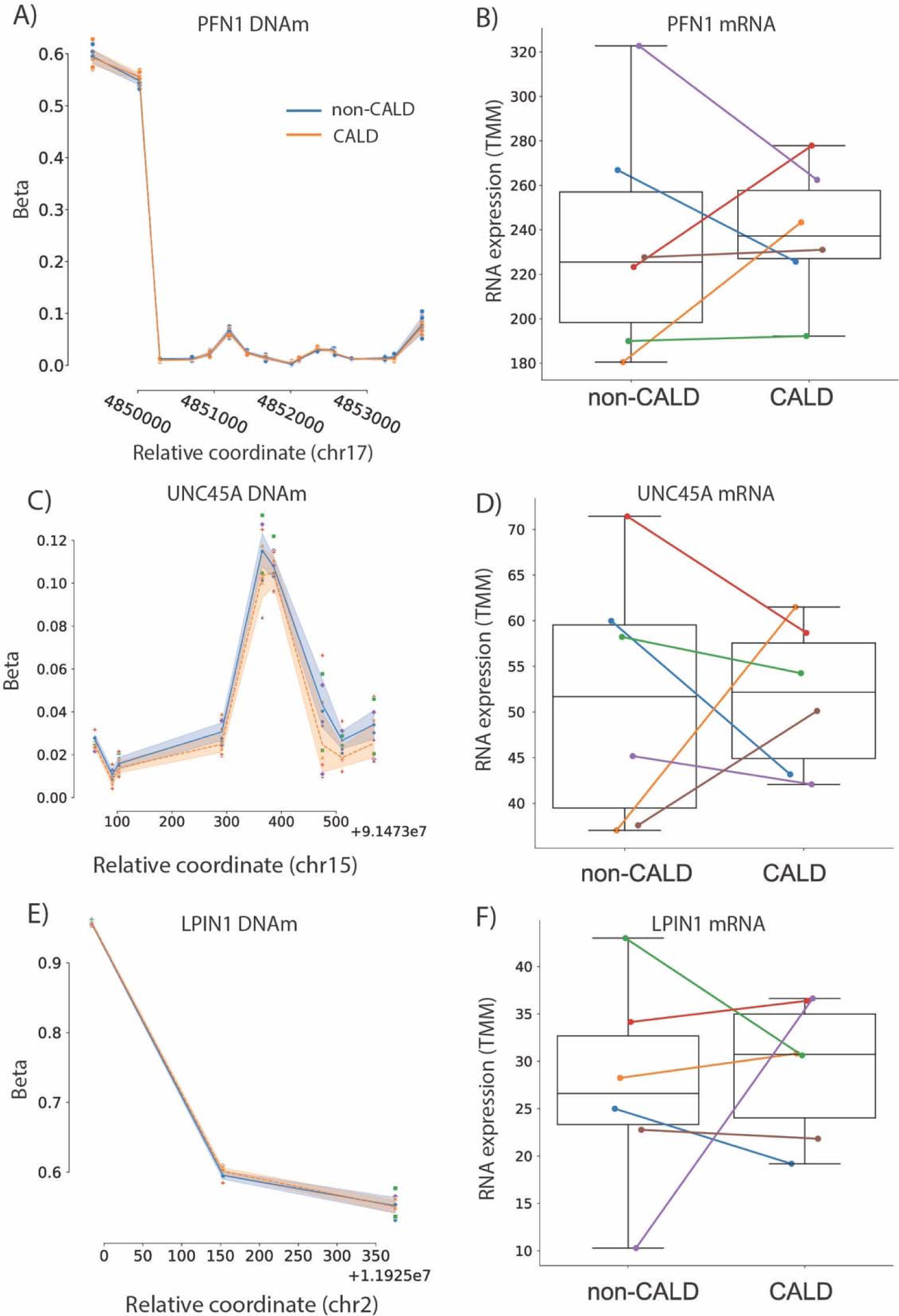
Testing previously suggested markers. DNA methylation and mRNA abundance for PFN1 (A,B), UNC45A (C,D), and LPIN1 (E,F). DNA methylation is shown for all CpGs associated to the listed genes, with the non-CALD mean methylation shown as a blue line with standard error shading, and the orange dashed line showing the mean methylation of CALD. Individual points are shown and colored by family. RNA expression is shown as a boxplot for non-CALD and CALD phenotype groups, with individual families labeled with family 1 as blue, family 2 as orange, family 3 as green, family 4 as red, family 5 as purple, and family 6 as brown.

Another study focused on DNA methylation (DNAm) as a marker of CALD, and investigated the intact white matter of brains from patients affected by ALD with and without the cerebral demyelination phenotype (Schlüter et al. 2018). Whether or not the signals they identify confirm within the blood within a separate cohort is important if these proposed marker genes are to be used within newborn screening. Within their analysis they identified differential methylation signals at several genes, two of which are *LPIN1* and *UNC45A*. Within this cohort, we see no differential methylation signal in the blood for LPIN1, and a slight hypermethylation (although not significant) in UNC45A (Figure 5C,E). Investigating the RNA shows that while both these genes are highly expressed, there are no consistent differences between the two phenotype groups.

Lastly, it is possible that a viral infection causing an immune response is the phenotypic trigger for progression to CALD, as this is suggested to be a candidate environmental modifier from other cerebral demyelination diseases including multiple sclerosis (Libbey, Lane, and Fujinami 2014). As is the case in several cancers, RNA-seq can capture actively expressing viral RNA within a sample. To test the hypothesis of whether or not we could observe different expressing viruses within the RNA-seq of these patients, we used the tool Centrifuge to identify traces of viral (or bacterial) sequences (Table S10) (Kim et al. 2016). Aside from identifying human, synthetic construct, and endogenous retrovirus, no significant viral or bacterial sequences were identified.

## 4 Discussion

In this study we took a systems biology approach to identify personal molecular characteristics, either genetic or molecular markers, which may prognosticate the onset of cerebral demyelination in patients affected by ALD. Identifying a single modifier consistent across all individuals has importance because of its potential utility as prognosticator or biomarker heralding the transition to cerebral demyelination, and this carries tremendous treatment implications.

Our cohort was comprised of carefully phenotyped brothers affected by ALD who were discordant for the severe cerebral demyelination phenotype. We collected blood and performed high throughput experiments to profile the DNA, methylated DNA, RNA, lipids, and proteins. In summary, we did not find a strong, convincing, univariate marker which can differentiate all of the CALD and non-cerebral patients in this small cohort. There are several explanations for this negative result: the small cohort with only six discordant sibling pairs of different ethnic background, the possibility that one or more non-CALD patients may still develop cerebral demyelination, high inter-individual variability, and finally the possibility of multiple modifiers and/or an exogenous or non genetic modifier such as infection or physical trauma. In spite of these limitations, we still emerged with interesting results from each of the omics platforms from this pilot study including discordant genotypes separating all CALD and non-CALD patients, confirmations of recently proposed CALD modifiers, and a suspected involvement of differential activity within the immune system in patients with cerebral demyelination.

In our genetic approach, we identified two discordant genotypes shared between all six brother-pairs: an intronic SNV in *DEUP1* and an SNV downstream of *WIBG*. Although in silico analysis of the variants and the function of the associated genes did not link these alleles to the cerebral demyelination phenotype, it is of interest to see if they replicate in a larger cohort.

Examining variants shared by all CALD patients led to the identification of a missense polymorphism in *TPCN2*, a gene which localizes to lysosomal membranes. This exists in a segregating haplotype block, and is absent from all non-CALD patients except for family 4--the youngest patient with the highest chance to develop cerebral demyelination--where the haploblock is homozygous. How this variant segregates in a larger patient cohort could be of interest. None of the previously proposed modifier alleles, emerging from GWAS or target-gene studies, confirmed within our cohort.

Although our analysis was burdened by high inter-individual variability, we were able to identify univariate molecular markers with increased confidence due to replication--by multiple omics levels and/or by confirming previously proposed modifier markers. A recent study (Schlüter et al. 2018) showed CALD patients with DNA hypermethylation within PTPRN2, which we confirm in our study and support with decreased mRNA expression in CALD patients (both platforms reaching p-value <0.05 before multiple testing correction). The same study showed hypermethylation of LPIN1 and UNC45A, the latter of which we confirm (although not statistically significant) as slightly hypermethylated in CALD samples. Of note, that study used brain tissue to derive their signal whereas we use blood samples. Another study utilized CSF and blood plasma, including longitudinal data from ALD patients pre- and post cerebral demyelination, to identify autoreactivity to Profilin 1 (PFN1) within CALD patients (Orchard, Nascene, et al. 2019). They observed auto-antigens to PFN1 in the blood, and increased PFN1 levels in CSF, in ∼50% of CALD patients. In our cohort, four out of six patients exhibit increased PFN1 protein levels, in-line with the observation that PFN1 phenotype is not ubiquitous across all CALD patients. We further contribute to this observation by showing no differences at the DNAm or mRNA levels, pointing towards a separate mechanism of upregulation/overabundance of PFN1.

As ALD is a peroxisomal disorder, the lipidomic analysis presented here is of interest. The lipid profiling data confirmed previous observations regarding VLCFA abundance differences in ALD samples when compared to controls. Specifically, the phosphatidylcholines (PC) species containing very long-chain fatty acids are more abundant in the ALD group compared to the control group. Furthermore, the suitability of LPC(C26:0) to function as a marker for ALD in newborn screening was confirmed. Differences in lipid abundance between CALD and non-CALD groups did not reach significance after multiple testing correction, likely due to a lack of consistent lipid differences between all brother pairs. Nevertheless, the differential lipids between CALD and non-CALD provide insight into the pathophysiology of CALD as CALD patients had lower levels of sphingomyelin and its precursor ceramide, in line with disease progression. This could support the findings that the sphingolipid systems hold important roles in CNS disorders like Alzheimer’s, Parkinson’s and Huntington’s (Assi et al. 2013).

Beyond identifying phenotype-stratifying molecular features, we investigated the top hits at the gene-level from each omics platform for any relation to the pathophysiology of CALD. Literature searches highlighted genes involved in lipid metabolism, the nervous system, and the immune system. Gene Ontology and KEGG pathways further supported these observations. Larger datasets are needed to draw conclusions from differentially abundant molecular features.

Throughout this work we have identified certain limitations of our approach which should be considered in future work focused on modifiers of rare disease, especially for other inborn errors of metabolism (e.g. Gaucher disease). First, we suffered from having a small number of samples and a high number of observed features. For future univariate marker investigations we recommend focusing only on protein or mRNA and increasing the number of samples. Second, our genetic analysis was limited by the possibility of future transition to the CALD state for any of our non-CALD patients, especially those patients who have not reached maturity. Recent epidemiological analysis shows that cerebral demyelination can occur throughout the lifetime of an ALD patient (Huffnagel, Laheji, et al. 2019), so genetic studies should focus on older (60-70 years old) patients who have not developed the cerebral demyelination phenotype. While discordant brother pairs reaching old age are challenging to find, a collection of genotyped non-CALD patients older than 60-70 years of age could serve as a good control. Third, we are limited in capturing relevant biological insights because we are profiling blood not CSF/brain tissue. Lastly, while we profile DNA methylation, we don’t capture other components of the environment which could have an impact including microbiome and pathogen exposure history.

With newborn screening now a reality for ALD, prognostication and timing of therapy becomes more relevant than ever before; thus modifier studies to decipher a protector or marker for cerebral demyelination will continue (Moser and Fatemi 2018). We believe that this dataset can continue to be mined and used for testing the replication of proposed phenotypic markers. Further, the data within this study could be used as part of a larger dataset examining multivariate signals differentiating the two classes. Whether it is a collection of genetic markers or a pattern of multiple molecular features, it is clear that there is a need for a larger sample size. As such, we make the measurements through this study available for future use to the community, with the hopes that the data can serve as a secondary confirmation of new modifier hypotheses, or as part of a larger dataset for investigating the complex nature of cerebral demyelination.

## Data Availability

The datasets generated and analyzed for this study can be found in the Zenodo repository: DOI- 10.5281/zenodo.3698292

https://zenodo.org/record/3698292#.XmrVHi0ZNTY

## 7 Author Contributions

Project was conceived by SK, ME, and CvK. Patients were recruited by ME. Paper written by PAR, FvdK, WW, CvK, SK, and ME. Data analysis for this work includes: WGS data analysis by PAR; RNA-seq analysis by FvdK; Lipidomics analysis by FV; Proteomics analysis by AU and PL; DNA methylation analysis by DL and MK; statistical analysis by EG, SM, PDM, and AHCvK.

## 9 Acknowledgments

We gratefully acknowledge the patients and families for their participation in this study, and the clinicians and colleagues for their expert management.

## 10 Data Availability Statement

The datasets generated and analyzed for this study can be found in the Zenodo repository: DOI-10.5281/zenodo.3698292; URL-https://zenodo.org/record/3698292#.XmrVHi0ZNTY.

Analysis code for this project can be found in the github repository: https://github.com/Phillip-a-richmond/ALD_Modifier_Project.

